# Time Series Modeling of Fatigue in the Stratification of Fatigue Phenotypes in Patients with Multiple Sclerosis

**DOI:** 10.1101/2025.05.16.25327776

**Authors:** Kezia Irene, Shayan Shokri, Alfredo M. Pinzon, Miklos Palotai, Charles R. G. Guttmann

## Abstract

Fatigue is a prominent paralyzing symptom that is often notified by patients with multiple sclerosis (MS). However, it is very difficult to measure fatigue because the symptoms fluctuate throughout the day. We analyzed the fatigue time series of 64 patients with chronic MS and found 6 distinct clusters that represent all patients: patients with low fatigue/ low MESOR (Midline Estimating Statistic Of Rhythm), lower-middle MESOR; positive acrophase, lower-middle MESOR; negative acrophase, middle-high MESOR; positive acrophase, middle-high MESOR; negative acrophase, and high MESOR. We also found that patients who are from the middle-high MESOR; negative acrophase cluster are more likely to be depressed compared to patients who are from the middle-high MESOR; positive acrophase cluster. The stratification of MS patients according to their fatigue phenotype and the difference in depression between different fatigue clusters could help future research on personalized MS treatments.

## 1. Introduction

Multiple Sclerosis (MS) is an incendiary and deteriorating chronic disease that affects the brain and spinal cord [1]. MS disrupts communication between different parts of the central nervous system (CNS), causing varied symptoms depending on the impacted circuitry. Symptoms can include numbness, blurred vision, bowel dysfunction, and cognitive impairment. According to Walton et al. [3], there are an estimated 2.8 million people worldwide suffering from MS. MS affects 2 out of 100,000 people in a year with females being more likely to suffer from MS.

Fatigue particularly is a major weakening symptoms of people suffering from MS [2], affecting around 50 to 90 percent of MS patients [12]. The pathogenesis of fatigue in MS patients is thought to be multi-factorial. Inflammatory mediators, metabolic factors, as well as neurodegeneration of specific brain circuitry (“central fatigue”) are thought to contribute to MS-related fatigue [6]. Many studies have shown an association between fatigue and changes in lesion, grey matter, white matter, and brain atrophy that characterize MS[2, 24, 25].

Fatigue is a subjective symptom, frequently assessed using standardized scales based on structured questionnaires, such as the modified fatigue impact scale (MFIS), the fatigue severity score (FSS), or a visual analog scale (VAS), leading to possible semantic ambiguity of the fatigue symptom between patients. The term fatigue can be interpreted as tiredness, muscular fatigue, cognitive fatigue, etc. The phenotypes of MS-related fatigue are various and difficult to be captured accurately using diagnostic surveys, such as MFIS or FSS. Many studies found that depression, anxiety, and physical activities are associated with MS-related fatigue [6,7]. Furthermore, fatigue is often not a constant symptom in MS. Rather, it can have a fluctuating temporal behavior. Taking comorbid symptoms as well as temporal behavior into account may help us define different fatigue phenotypes, which in turn might reflect different pathogenic mechanisms. By stratifying MS patients according to different fatigue phenotypes, we might explore each group’s mechanism and, eventually, seek more personalized treatments for fatigue, which are currently lacking.

Previous work by Shokri et al. (unpublished) in Dr. Guttmann’s group aimed at identifying robust and distinct fatigue phenotypes in MS patients, by various statistical and unsupervised clustering methods applied to VAS of fatigue, anxiety, depression, and pain. Preliminary findings of this work indicate that fatigue, but not anxiety, depression, or pain, shows significant circadian variation. Furthermore, patients with moderate-to-high levels of fatigue can be further stratified according to the presence or absence of significant depression. Shokri et al. also proposed the stratification of MS patients into 6 different clusters based on a decision tree approach applied to the measured VAS and fatigue VAS temporal behavior.

Currently, the disease development of fatigue in MS is not well understood and there is a lack of effective treatment for this debilitating symptom. Campo, CG et al. [9] developed a machine learning model to determine if MS-related fatigue is associated with changes in the behavior, structure, and function of the brain using a one-time fatigue impact scale. Tong et al. [10] proposed a machine-learning approach to predict fatigue scores from MS patients that were using connected wellness devices for 6 months, achieving a mean absolute error of 1.00 on a scale with 7 discrete levels. However, none of these studies utilize the time-dependent nature of fatigue in MS patients throughout the day.

Therefore, our research group developed an online application to explore fatigue behavior using diurnal monitoring (every 4 hours during waking time) of fatigue, and other symptoms such as depression, anxiety, and pain [8]. Using this detailed description of fatigue and other symptoms that resulted from MS, we propose to stratify MS patients based on these attributes (fatigue visual analog scale, depression analog scale, anxiety analog scale, and pain analog scale) and then find out how the fatigue phenotypes in each group associate with the change in the anatomical structure of the brain.

The primary objective of this study is to determine whether different fatigue phenotypes reflect distinct patterns of neurodegeneration affecting different brain circuits. To achieve this, we stratified and test the robustness of our method to determine the fatigue phenotypes in MS, using time series approaches. We also did some preliminary development to assess whether different fatigue phenotypes are reflected in different brain MRI signatures, using grey matter and white matter parcellation techniques.

This aim of this study is to enable physicians to differentially assess specific treatments of fatigue in groups of MS patients with different fatigue phenotypes. Understanding patient stratification could provide MRI predictors of treatment response to help clinical decision-making by selecting the best fatigue management intervention for individual patients.

## 2. Patients and Methods

### 2.1 Study Population

We recruited 2400 patients suffering from MS for the Comprehensive Longitudinal Investigation of Multiple Sclerosis at Brigham and Women’s Hospital (CLIMB) study [8]. CLIMB is a large-scale study for. The patients were recruited using the following criteria: (1) Having a record of the Modified Fatigue Impact Scale (MFIS) (2) Having a brain MRI scan within a month of study, and (3) Having no worsening within 3 months of brain MRI acquisition [8]. We proposed to study deeper about fatigue in daily life with an additional study with the same patient population to learn more if fatigue symptoms change throughout the day.

From 2400 patients, we recruited a subset of 64 patients and invited them to an in-person study initiation session (SIS) at the Brigham and Women’s Hospital. During the SIS, the patients were informed of the detailed study description, signed a consent form, and were taught to utilize an in-house built mobile app [8], a wrist actigraphy watch [26], and a sleeping test device [27].

Using the in-house built mobile app [8, we monitored the visual analog scale (VAS) of fatigue, depression, anxiety, and pain for every patients. The application will ask the patients to fill the VAS every 4 hours (except the patient labeled themselves as asleep). The actigraphy watch monitored patients’ activity and sleep quality. Finally, the sleeping test device measured patients’ sleep apnea and movement during their sleep.

Table 1 presents the demographic data of all 64 patients. Each patient was awarded a sum of $100 for their contribution.

**Table 1.** Demographic information and patients completion record [8].

| Variable | All Patients | Group 1: Completed all one-time questionnaires, VAS, and SLD | Group 2: Completed all one-time questionnaires, some VAS and SLD | Group 3: Did not complete one-time questionnaires, VAS, or SLD |
| --- | --- | --- | --- | --- |
| Subjects, n (%) | 64 (100) | 47 (73) | 4 (6) | 13 (21) |
| Age, mean (SD) | 52 (9) | 52 (8) | 54.4 (10) | 51 (9) |
| Sex (female), n (%) | 54 (84) | 40 (85) | 4 (100) | 9 (70) |
| White (%) | 61 (95) | 45 (96) | 4 (100) | 12 (92) |
| Disease duration (years), mean (SD) | 20 (8) | 21 (8) | 28 (10) | 17 (7) |
| Fatigue Severity Scale score, mean (SD) | 42 (13) | 42 (14) | 47 (12) | N/A |

Note: Adapted from Palotai et al. (2021) [8]

### 2.2 Patient Compliance

We recruited 64 patients, however, 8 patients (13%) never answered any questionairre or record any sleeping diaries in the mobile application. 5 patients (8%) never completed the visual analog scale and 3 patients (5%) completed it in less than 8 days, therefore we are unable to include these patients in our study because of the lack of data. Figure 1 shows patient compliance in our study cohort.

**Figure 1.**
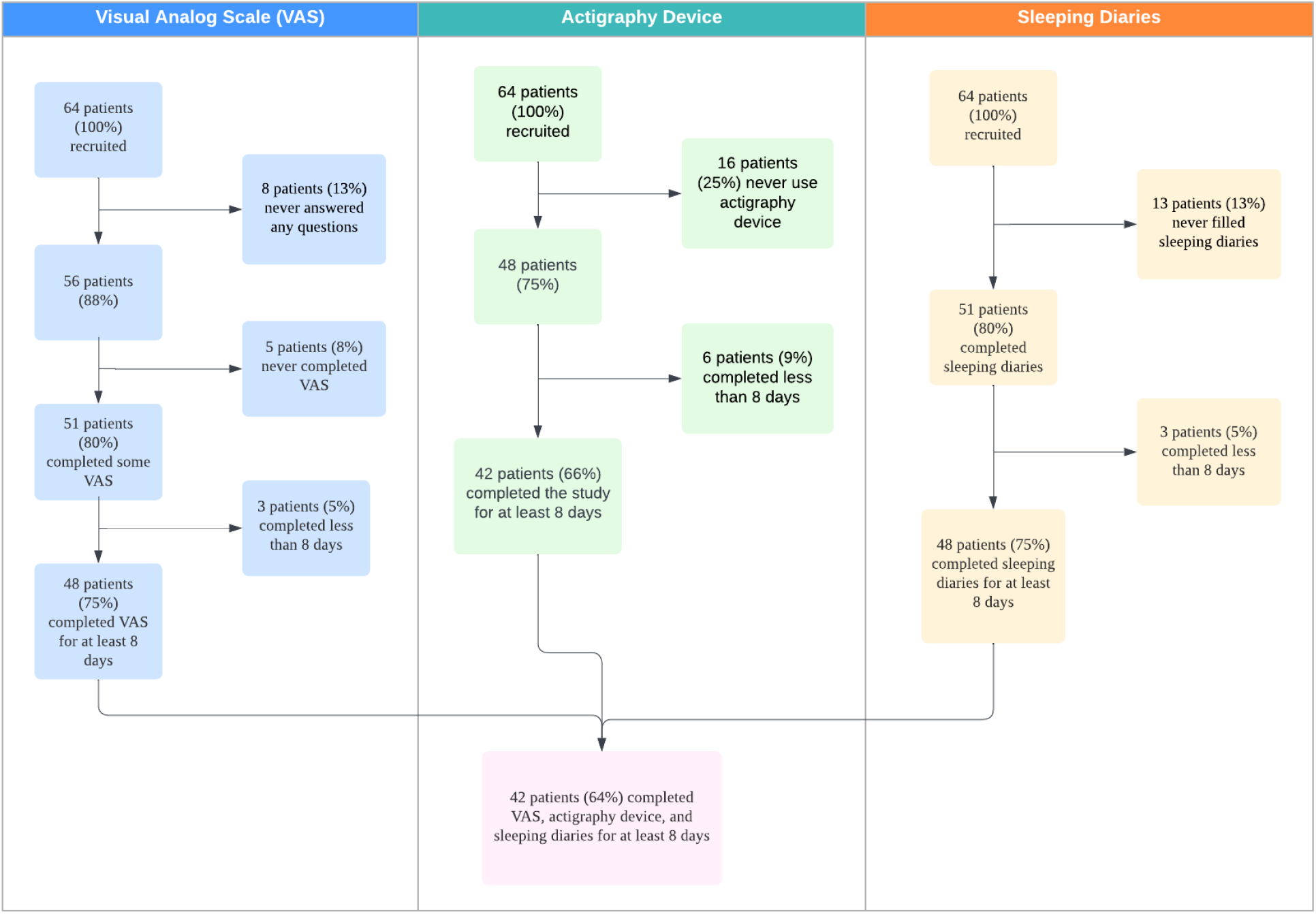
Patient compliance in our study cohort.

16 patients (25%) never use any actigraphy device. Furthermore, 6 patients (9%) utilized the actigraphy device for less than 8 days. Therefore, the remaining 42 patients (66%) had at least 8 days of data of visual analog scale answers, actigraphy device use, and sleeping diaries. We never received any reported issues regarding the mobile app and the data collecting system.

### 2.3 Study Design

We wanted to study the fatigue phenotypes in MS patients by exposing the circadian rhythm to cluster patients into different clusters according to their fatigue phenotype. The methods can be seen in Figure 2. First, we selected the eligible patients and standardize the data, then we remove the outlier days within individual patient days. Next, we used the cosinor function to interpolate each patient’s typical days. We applied a dynamic time-warping algorithm to find the similarities between different patient time series and applied hierarchical clustering to group patients’ fatigue phenotypes. We also used the cosinor parameters which are mesor (mid-line of cosinor wave), acrophase (the time from the lowest peak to the highest peak), and amplitude (half of the variation in a cycle)[5] to make a separate clustering using K-means. Finally, we combine the hierarchical and K-means clustering to make robust clusters.

**Figure 2.**
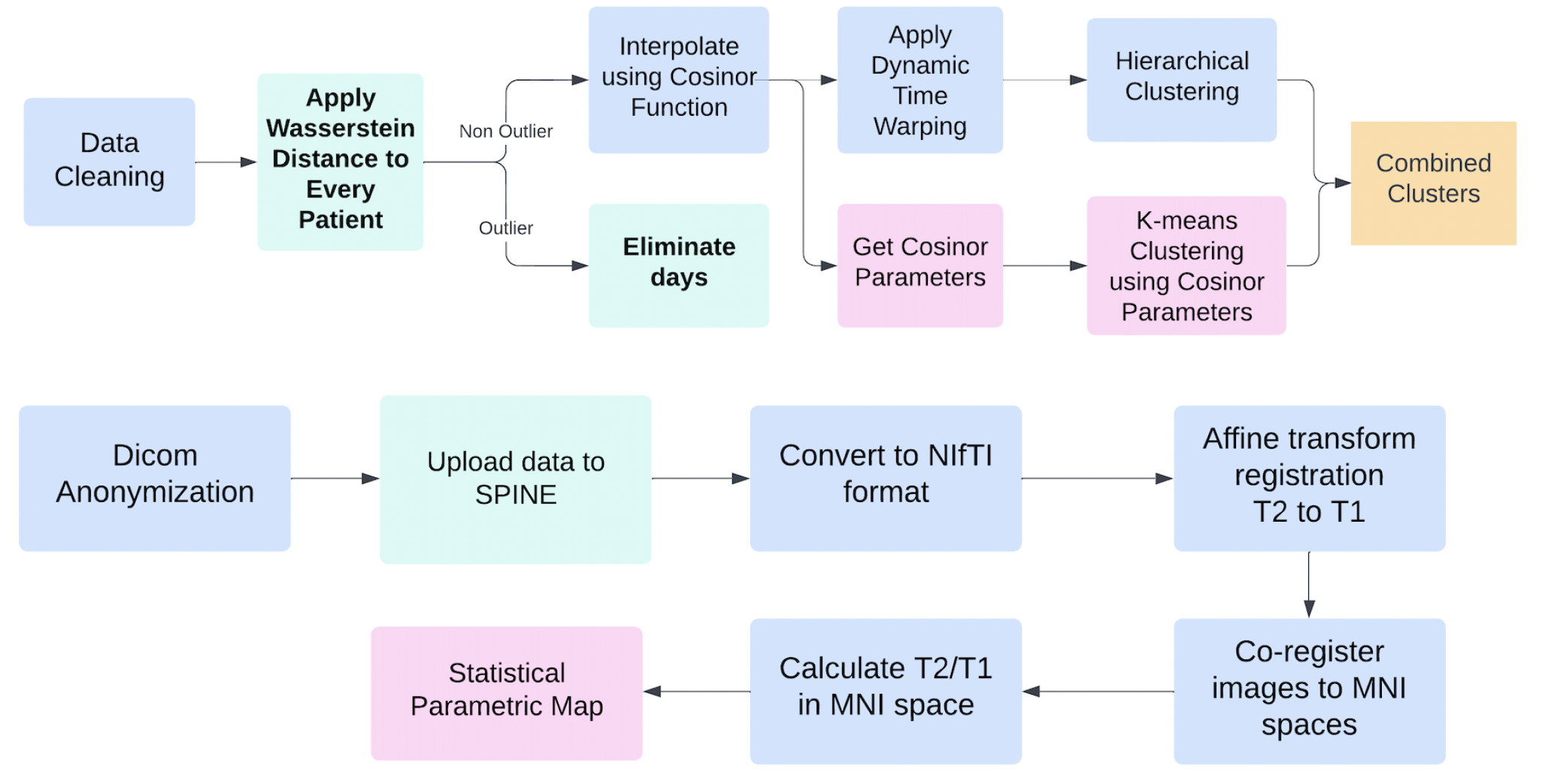
Methods flowchart. Top: fatigue phenotype clustering method, Bottom: brain MRI images processing method

We processed MRI images first by anonymizing Dicom metadata to protect patients’ personalized health information (PHI), then we uploaded the anonymized data to SPINE (Structured Planning and Implementation of New Explorations) platform [13]. After that, we plan to convert the images to NIfTI format because many of the modules in SPINE are implemented using NIfTI format. Then, we plan to co-register the images, apply T1/T2 weighted images to the MRI, move the MRI to standardized MNI spaces, and finally compute the statistical parametric map of each patient’s MRI. We have not finished the last 5 steps of the brain MRI image processing, thus the result is not mentioned in the result section.

#### 2.3.1 Selecting Eligible Patient Data

In the mobile application, patients answered visual analog scales (VAS) from a range of 0 to 10 to report fatigue, depression, anxiety, and pain levels every four hours with 2 hours tolerance to complete filling the VAS [8]. Patients have 2 sleep-awake monitoring devices: a mobile application and an actigraphy device [8].

There are a lot of granularities between patients and even individuals on different days. Patient changes their sleeping schedule every day and they also fill the VAS at a different time each day and there is some variation in the length of days patient filling. To normalize, we use hours from wake-up time as the measurement of a patient. We have 2 devices that monitor wake-up time, which are sleeping diaries and an actigraphy device. Figure 3 shows our data cleaning pipeline to normalize every single patient day.

**Figure 3.**
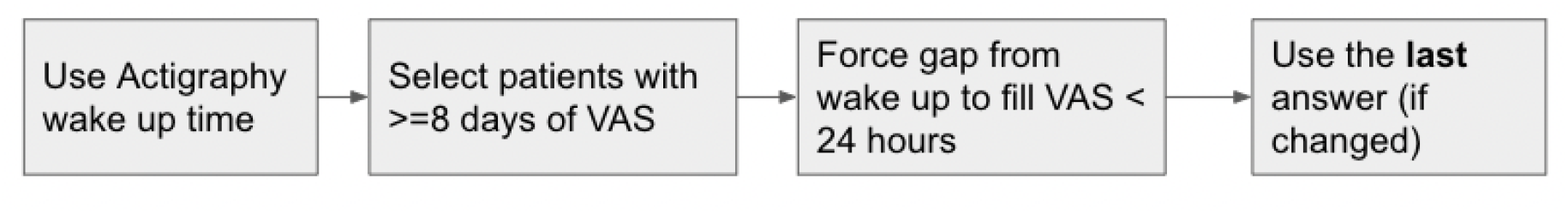
Data Cleaning Pipeline.

We compared sleeping diaries and actigraphy device data to select which sleeping device monitoring data we should use. For sleeping diaries, we found that 47 out of 64 patients have at least 1 missing day. For the actigraphy device, we found that only 15 out of 64 patients have at least 1 missing day. We also found that patients who have missing days in the actigraphy device also did not answer most of the VAS, therefore they have been eliminated in the first data cleaning pipeline.

We also found that the time gap between the nearest waking up time and filling the VAS is more reasonable. Most of the gap between waking up and filling VAS in an actigraphy device is less than 24 hours. Therefore we chose actigraphy wake-up time

#### 2.3.2 Outlier Day Elimination

We suspected that during 13 days of monitoring, patients might have outlier days where their fatigue VAS is different from other days. We computed the optimal transport solution using the Wasserstein metric [4] to measure the difference in distribution between days in an individual patient.

First, we interpolate the patient data using cosinor interpolation. Then, we computed the Wasserstein distance for every combination (day 1-day 2, day 1-day 3, day 2 - day 3, etc.). We defined a day to be an outlier if the mean distance between one particular day compared to any other day is larger than 1.5 IQR lower than the first or higher than the third quartile. We eliminated these days so we could get a “typical day” for each patient.

#### 2.3.3 Interpolation for Fatigue Time Series

Every eligible patient on average answered the VAS 4 times per day at a different time during the day. To process the data as time series, we combined every patient day that we have for one patient and then tried different interpolation methods that best capture the variance of our data.

According to Shokri, et al. (unpublished), fatigue in MS patients has shown a circadian pattern that is similar per day. To capture the circadian rhythm, we interpolated our data with the Cosinor function [5]. Cosinor-based rhythmometry is an interpolation procedure that tries to capture the nature-like circadian rhythm within a period of time, usually a day, a week, or a month [5]. Circadian rhythm has been applied for interpolating many biological circadian rhythms such as melatonin variation [11], cortisol regulation [18], and other endogenous rhythms [19,20].

Figure 4 shows the definition of acrophase, amplitude, and MESOR (Midline Estimating Statistic Of Rhythm)[5]. MESOR is the adjusted average of patients’ fatigue VAS answer. Amplitude is the distance between MESOR and the highest peak of the cycle. Acrophase is the measure of timing between the lowest valley (lowest fatigue) to the highest peak (highest fatigue). A positive acrophase means that the patient is less fatigue during the time after they wake up and more fatigue hours after they wake up. A negative acrophase means that the patient is more fatigue when they wake up and becomes less fatigue hours after they wake up.

**Figure 4.**
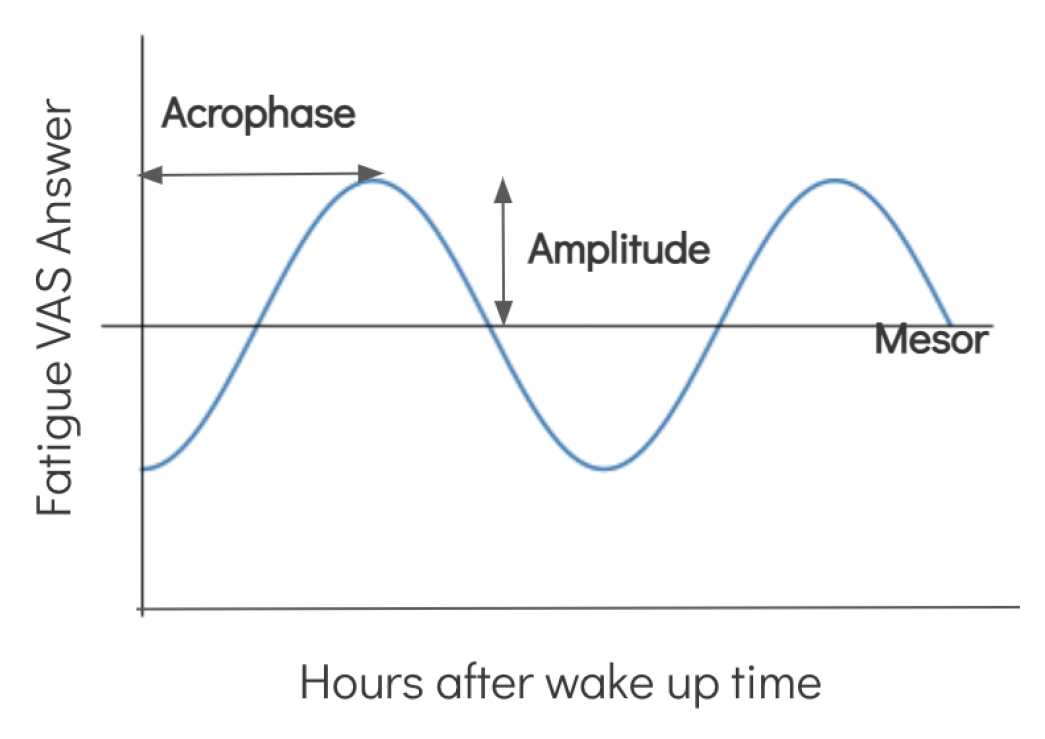
Diagram showing acrophase, aptitude, and MESOR (Midline Estimating Statistic Of Rhythm) in cosinor interpolation.

To measure the appropriate interpolation method, we compare the cosinor interpolation and linear interpolation to see which interpolation method captures the most variance within our data.

#### 2.3.4 Unsupervised Clustering

To capture the similarities between each patient’s time series, we use a dynamic time-warping algorithm. Dynamic time warping calculates the best match between two different time series where the algorithm match one time series to another time series with one extra or one minus index [21]. After getting the similarity matrix, we computed the cluster using hierarchical clustering [22]. Hierarchical clustering builds a dendrogram that places the time series that are similar the closest to each other.

We applied K-means clustering to get more information just using the cosinor interpolation parameters. K-means algorithm will partition our patients based on mesor (mid-line of cosinor wave), acrophase (the time from the lowest peak to the highest peak), and amplitude (half of the variation in a cycle)[5], into K groups that have the lowest distance to the center of the cluster [23].

#### 2.3.5 Statistical Parametric Map Analysis

Currently, we have anonymized patient data, uploaded the anonymized data into SPINE, and created a pre-processing script in JSON format. We listed the plan we want to do with the MRI image data.

##### 2.3.5.1 Dicom Patient ID Anonymization

We implemented a python script to anonymize patient ID given the input folder path and lookup table. We want to be able to map a patient to their relevant phenotype dataset. We also anonymized other metadata information that contains personal health information (PHI).

##### 2.3.5.2 Pre-Processing

The pre-processing will be done using SPINE. We plan to convert the images to NIfTI format because many of the modules in SPINE are implemented using NIfTI format. Then, we plan to co-register the images, apply T1/T2 weighted images to the MRI, move the MRI to standardized MNI spaces, and finally, compute the statistical parametric map of each patient’s MRI.

Figure 5 shows the example of our MS patient data. In T2-weighted and T2 flair-weighted images, we can see that this patient has a suppressed ventricle and sulcide (the surface of the brain is folded, and the sulcus is the valley in the fold) of the brain. We will normalize the signal by dividing T2-weighted images by T1-weighted images to make our images more stable across the different patients who might have different calibrations.

**Figure 5.**
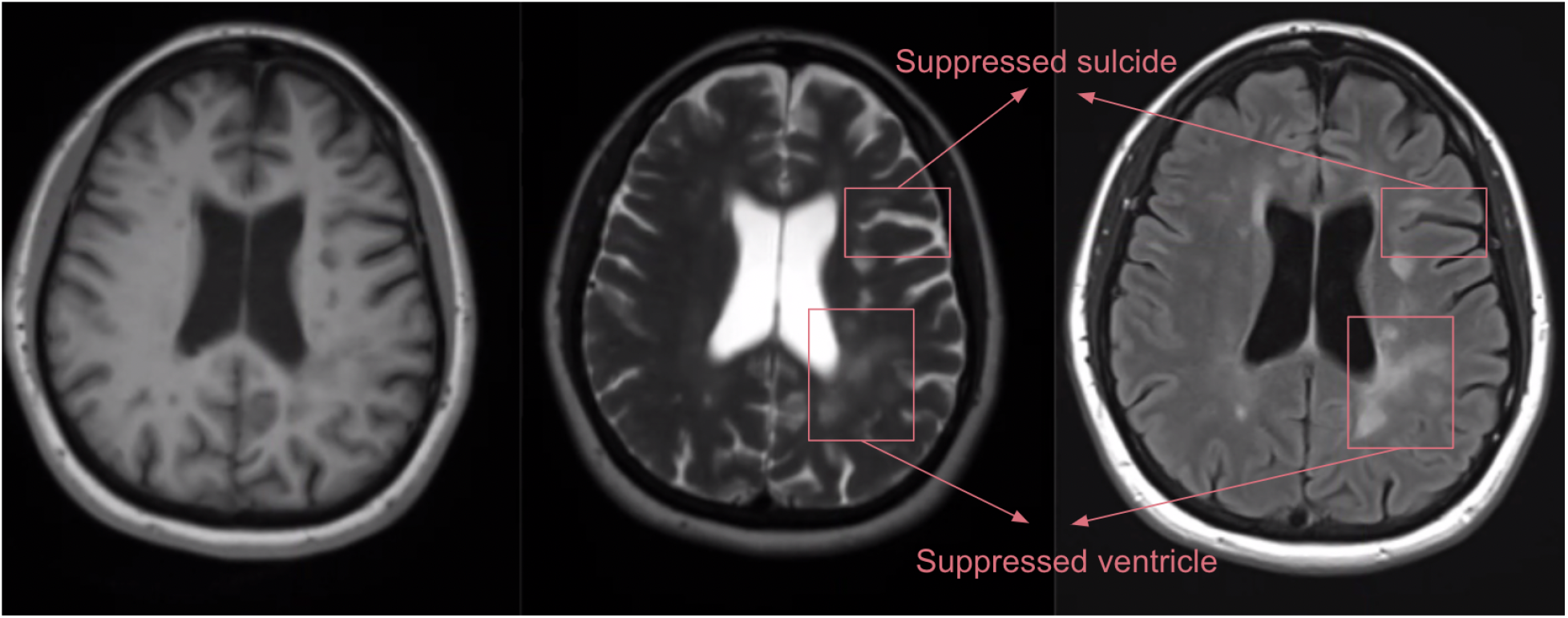
Multiple Sclerosis Patient MRI sample. Left: T1 weighted image. Middle: T2 weighted image. Right: T2-flair weighted image

## 3. Results

### 3.1 Eliminated days

Using the Wasserstein metrics combination and some manual cleaning, we eliminated outlier days for every patient. Figure 6 shows similar days within one patient, while the last picture shows a very different day. We eliminated this day because we consider this day as an outlier day. We found that most of the eliminated days were either Saturday or Sunday, indicating that there might be some different pattern of fatigue during the weekends.

**Figure 6.**
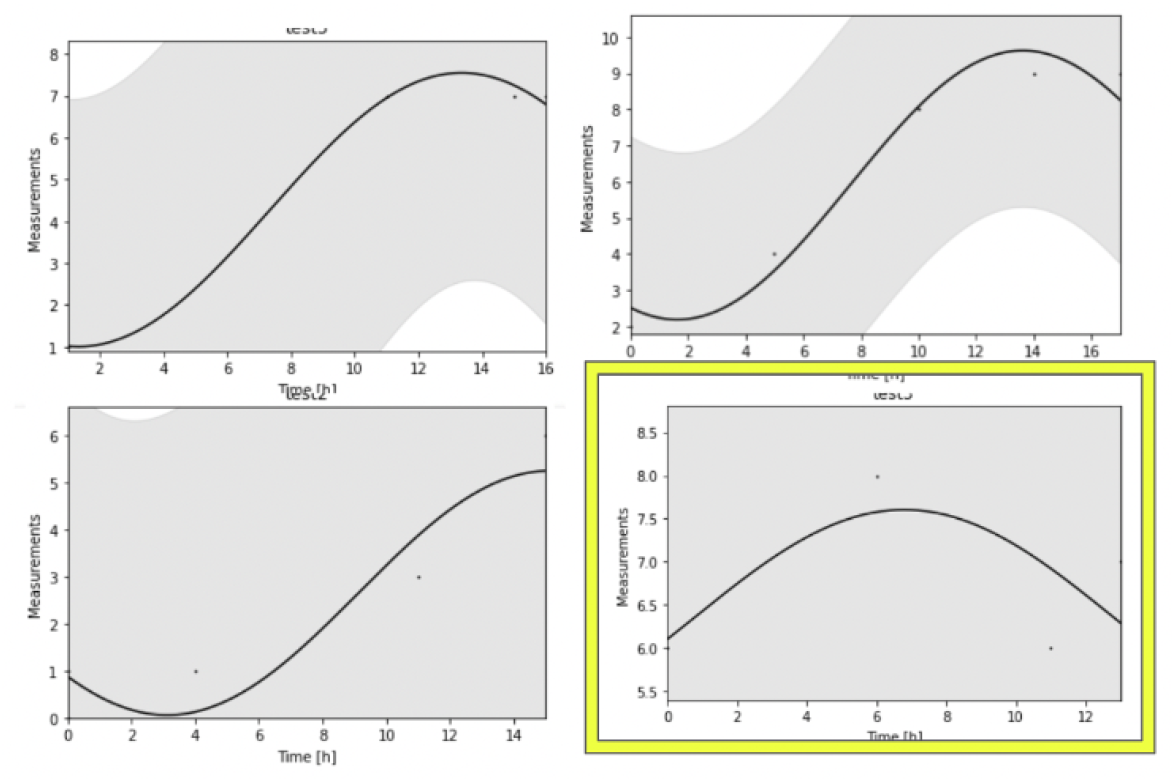
Example of an outlier day for a patient.

### 3.2 Cosinor interpolation captures more information about patients’ circadian rhythm

We compared the adjusted R2 score and the residual sum of squares (RSS) between cosinor interpolation and linear interpolation to see which interpolation method fit better to our data. In Figure 7, we found that in almost all cases, the R2 score for cosinor interpolation is higher than the R2 score for linear interpolation, meaning that cosinor interpolation is able to explain more variation in our data compared to linear interpolation. We also found that for most of the patients, cosinor interpolation has lower RSS meaning cosinor interpolation has a better fit compared to linear interpolation.

**Figure 7.**
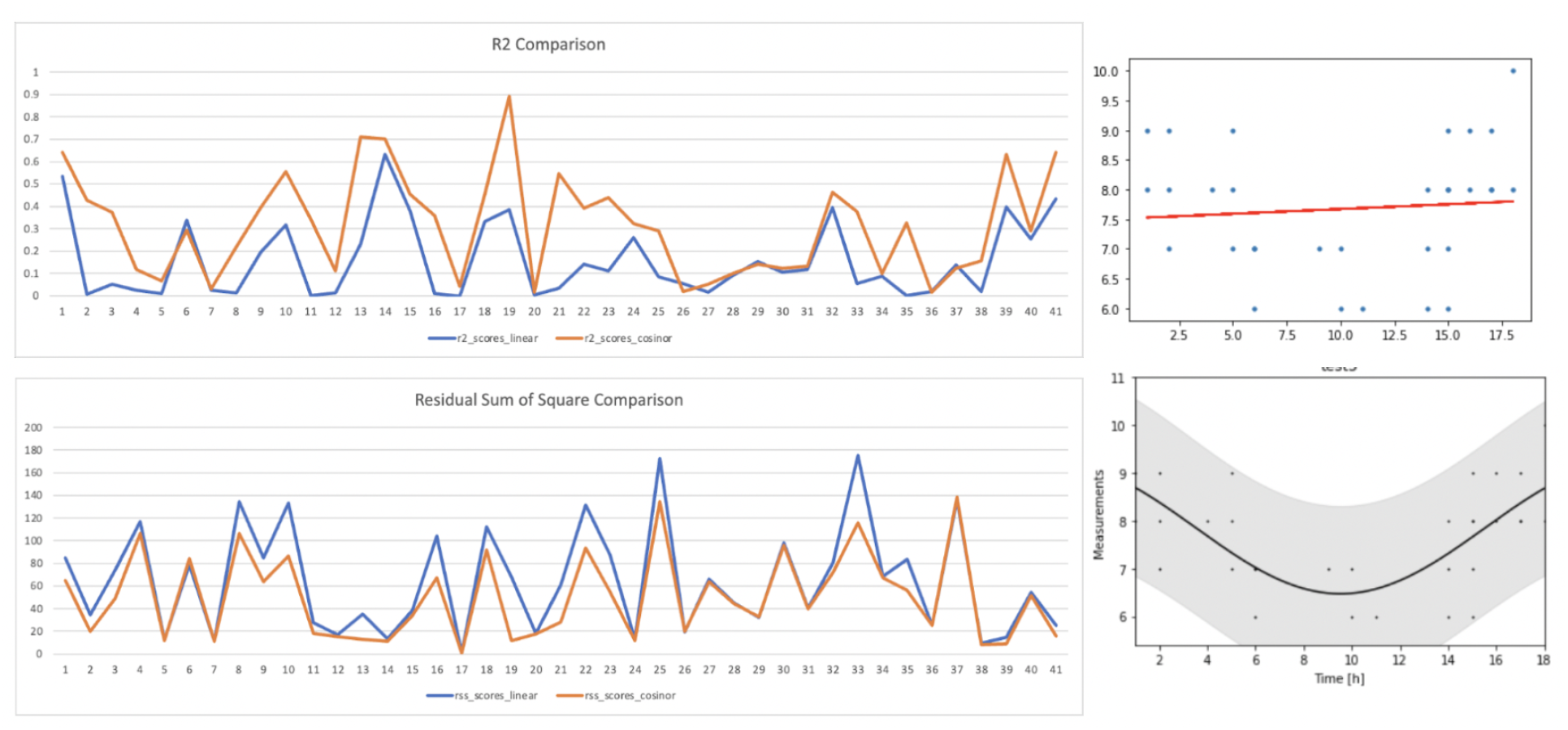
**Left:** R2 and residual sum of square comparison between linear interpolation (blue) and cosinor interpolation (orange). Top right: example of linear interpolation. Bottom right: example of cosinor interpolation

### 3.3 Clustering Result

Using dynamic time warping (DTW) and hierarchical clustering, we found 4 distinct clusters. We utilized agglomerative clustering with each dynamic time-warping distance as the distance function. We used a single-link linkage criterion and set 50 as the distance threshold to cut the tree. Our cluster dendrogram can be seen in Figure 8.

**Figure 8.**
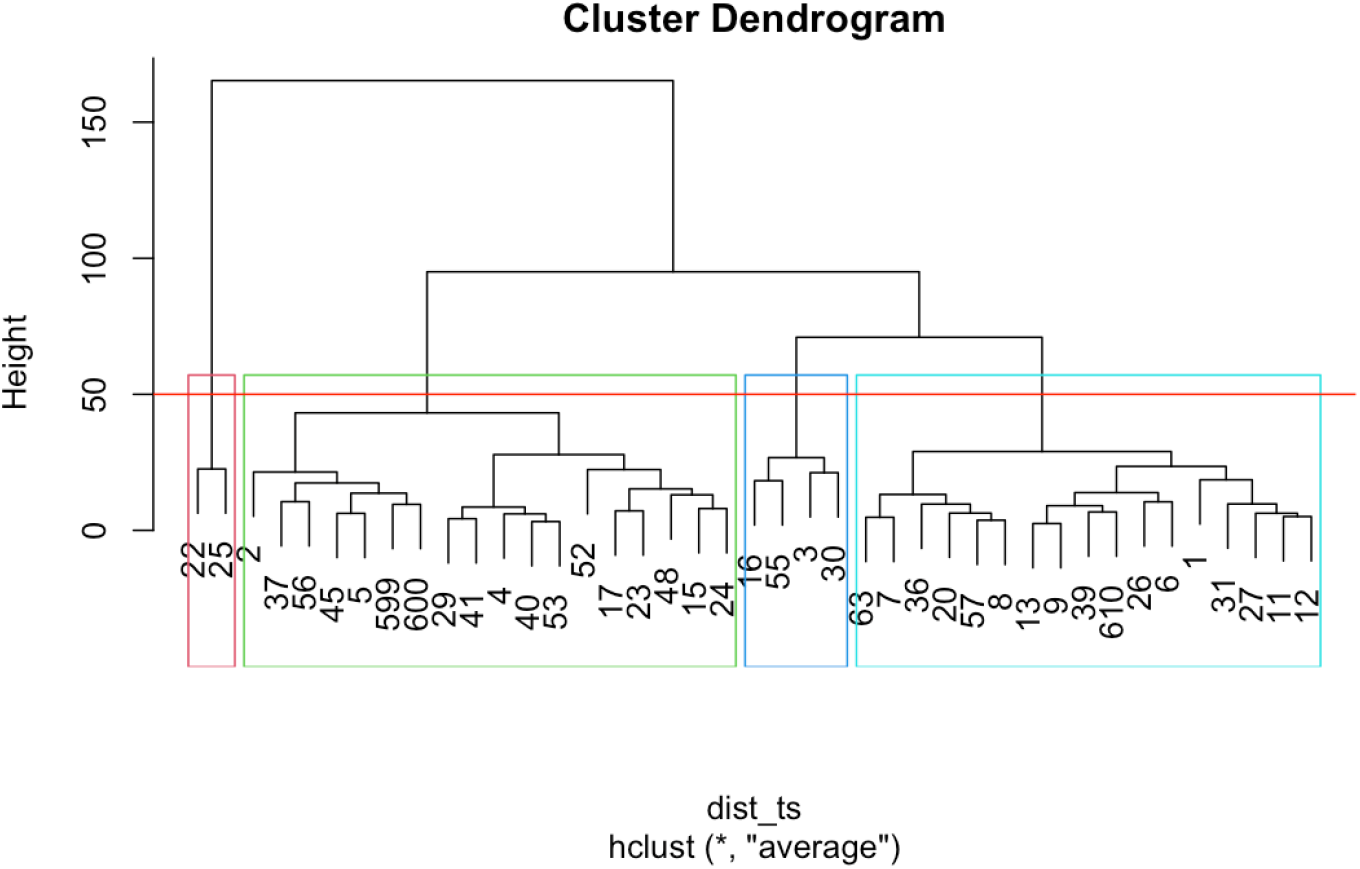
Hierarchical clustering with dynamic time warping results.

We then compare the result with K-means clustering using the parameter of cosinor interpolation, which are mesor, acrophase, and amplitude. We selected 4 clusters based on three classification performance indexes, which are the Calinski-Harabasz score, silhouette score, and Davies-Bouldin score. The result can be seen in Table 2.

**Table 2.** K-means classification performance analysis.

| Number of k | Calsinski Scores | Silhouette Score | Davies-Bouldin Score |
| --- | --- | --- | --- |
| 2 | 32.40 | 0.43 | 0.99 |
| 3 | 35.52 | 0.39 | 0.87 |
| 4 | **34.36** | **0.38** | **0.84** |
| 5 | 34.82 | 0.33 | 0.86 |
| 6 | 35.01 | 0.33 | 0.89 |
| 7 | 36.82 | 0.36 | 0.79 |
| 8 | 40.20 | 0.38 | 0.76 |

The hierarchical clustering based on dynamic time warping result mostly split the cluster based on mesor, while the K-means clustering result mostly split the cluster based on mesor and acrophase. We combined this information from 2 different clustering methods to get a combined clustering result in Figure 9. Then, we created a decision tree to find the exact range for mesor and acrophase (Figure 10).

**Figure 9.**
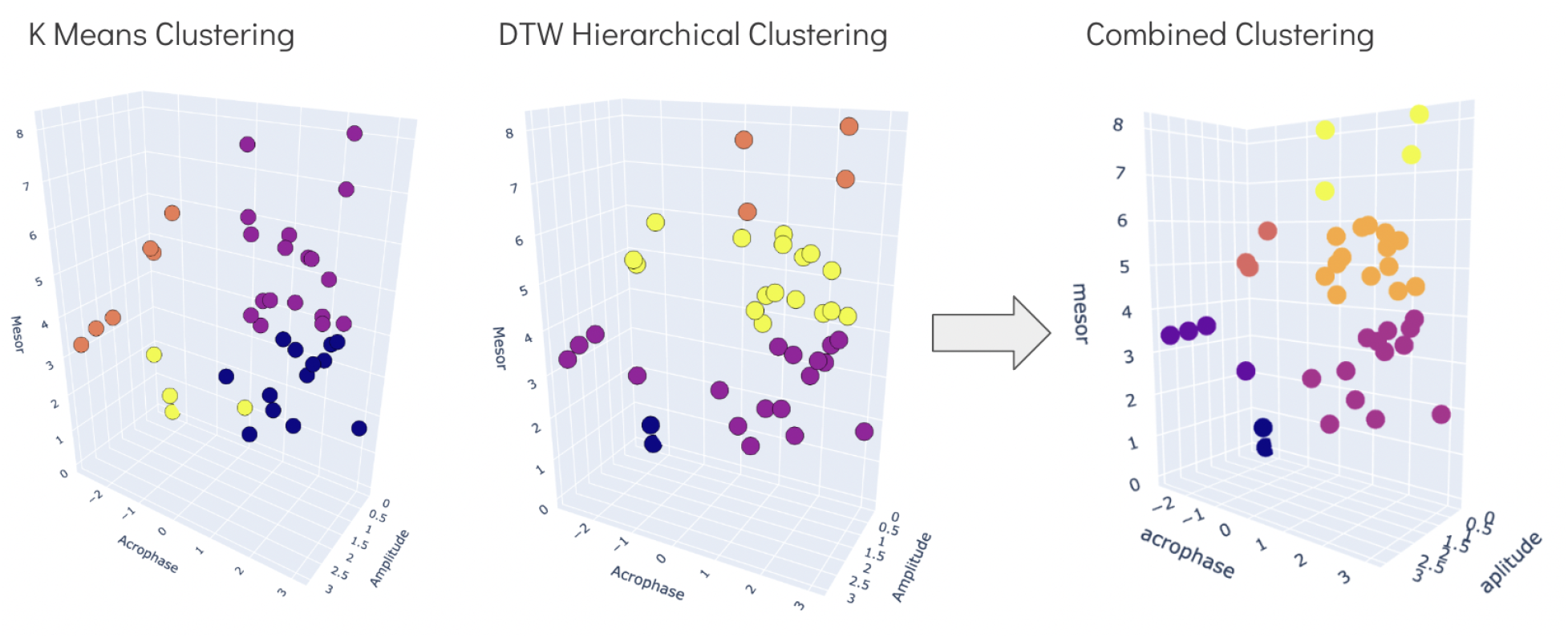
Combining results from K-means clustering and dynamic time-warping hierarchical clustering.

**Figure 10.**
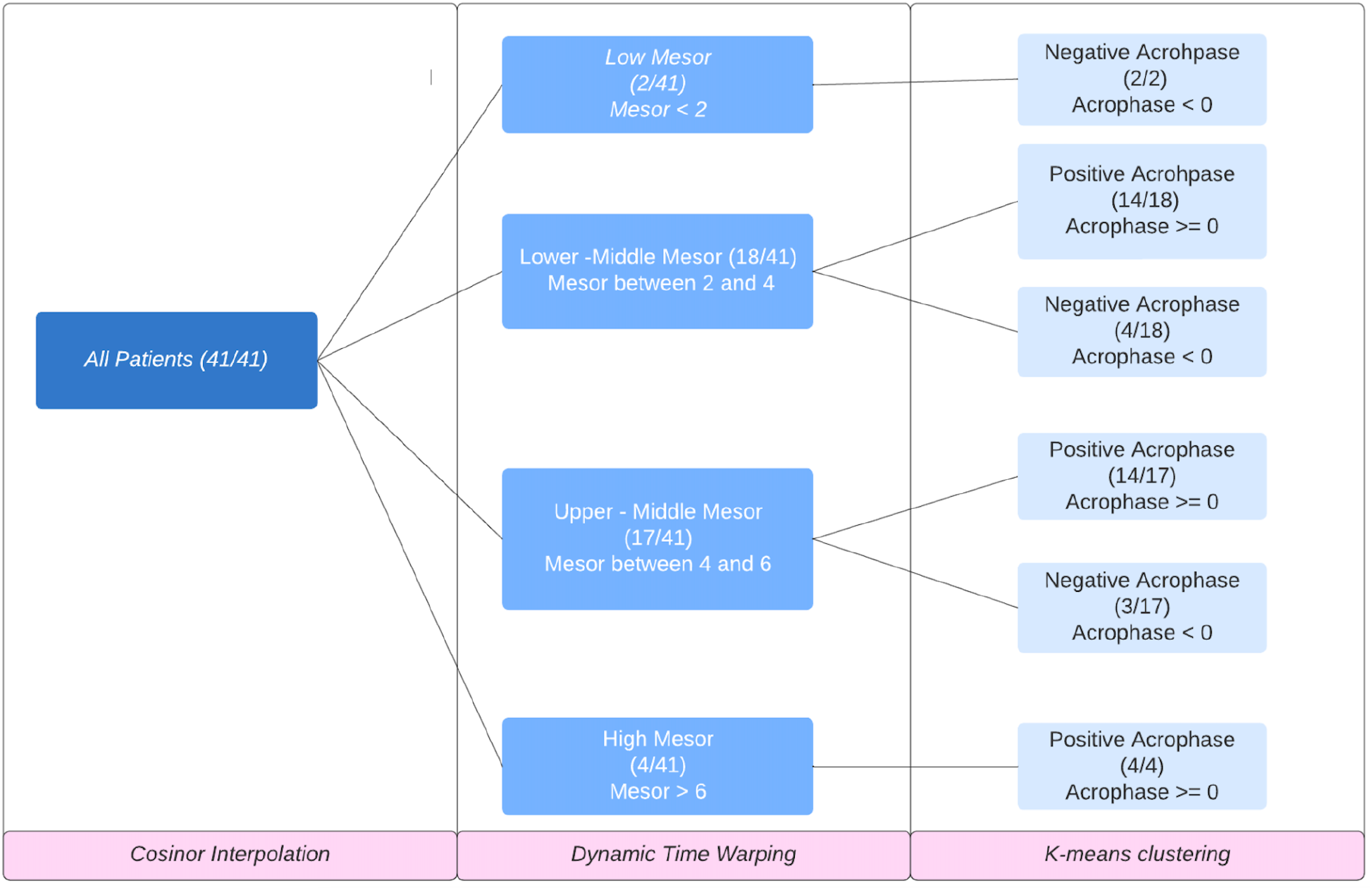
Summary of the decision tree for the total recruited MS patients.

From figure 10, we can see that most of the patients fall into the lower-middle mesor; positive acrophase cluster and upper-middle mesor; positive acrophase cluster, indicating that many patients feel more fatigue when they wake up compared to hours after they wake up. This could indicate that MS could possibly contribute to a lower quality of sleep.

Figure 11 shows the example of patients from each cluster. The first cluster contains 2 patients who are never fatigued, and have low mesor (<2) and negative acrophase (<0). The second cluster contains 14 patients with lower middle mesor (between 2 and 4) and positive acrophase (>=0), meaning these patients are more fatigue during the later hour after waking up compared to when they first wake up. The third cluster contains 4 patients with similar mesor to the second cluster, but negative acrophase (<0), meaning these patients are more fatigue during the time they wake up compared to hours after that. The fourth cluster contains 3 patients with upper middle mesor (between 4 and 6) and positive acrophase. The fifth cluster contains 13 patients with similar mesor to the fourth cluster and negative acrophase. Finally, the sixth cluster contains patients with high fatigue, as indicated by high mesor, and positive acrophase.

**Figure 11.**
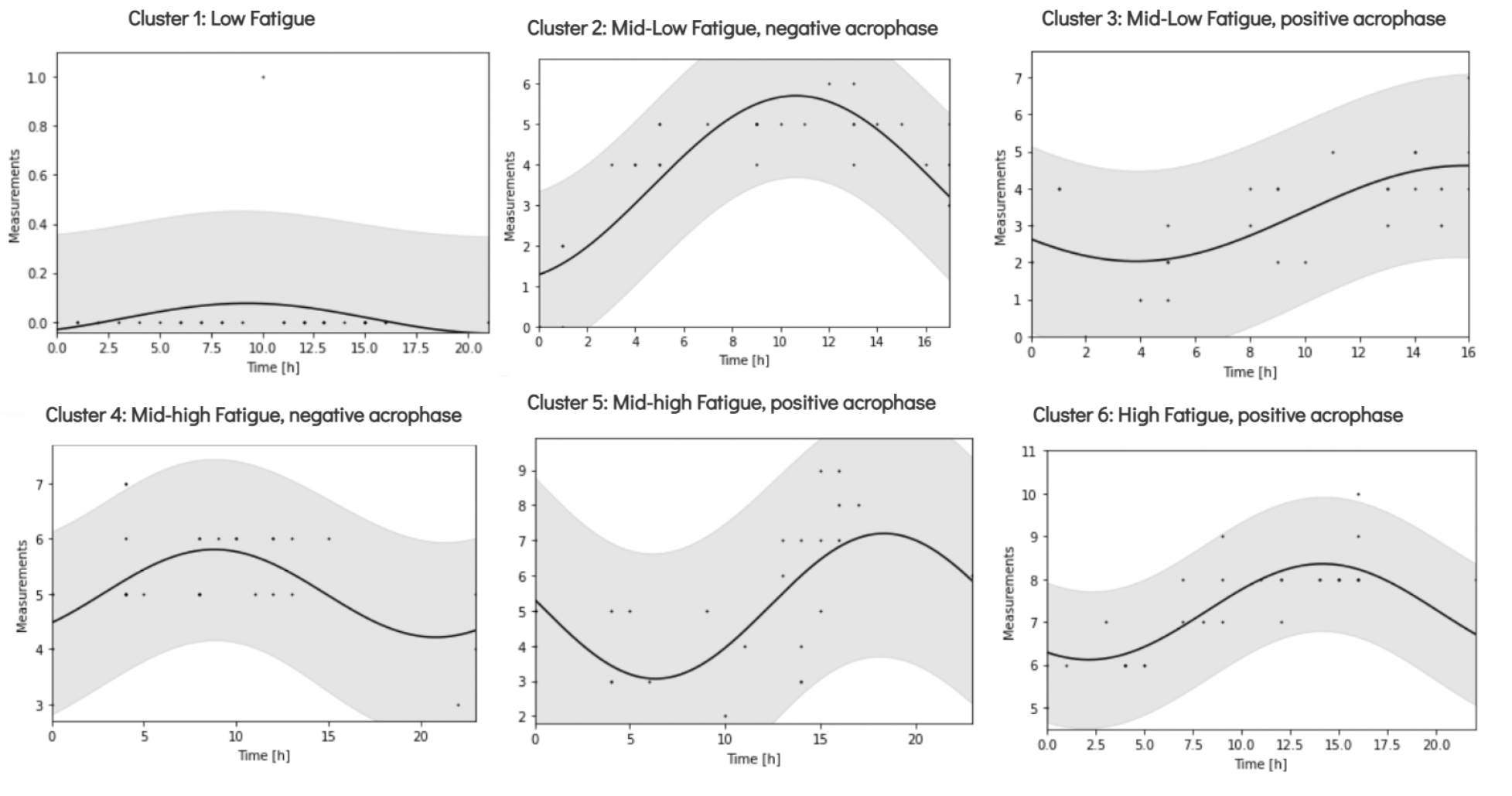
Cosinor interpolation for patients in each cluster. Top left: Low fatigue; negative acrophase. Top middle: Lower-middle mesor; negative acrophase. Top right: Lower-middle mesor; positive acrophase. Bottom left: Middle-high mesor; negative acrophase. Bottom middle: Middle-high mesor; positive acrophase. Bottom right: High mesor, positive acrophase.

### 3.4 Relation between fatigue cluster and depression

We also want to find the relation between our fatigue cluster and depression to see if depression is different within each cluster. First, we find the overall distribution of depression and decided to make 3 clusters, which are patients with no depression (median answer 0), patients with slight depression (median answer between 1 and 3), and patients with depression (the median answer is more than 3). For each cluster, we mapped the depression level and found that patients in the low fatigue and low-middle mesor with positive acrophase cluster are not depressed, and half of the patients in the lower-middle mesor with negative acrophase are slightly depressed. Most of the patients who are in the middle-high mesor are slightly depressed. However, in the middle-high mesor and negative acrophase cluster, all of the patients reported depression, although their mesor is similar to the previous cluster (Figure 12).

**Figure 12.**
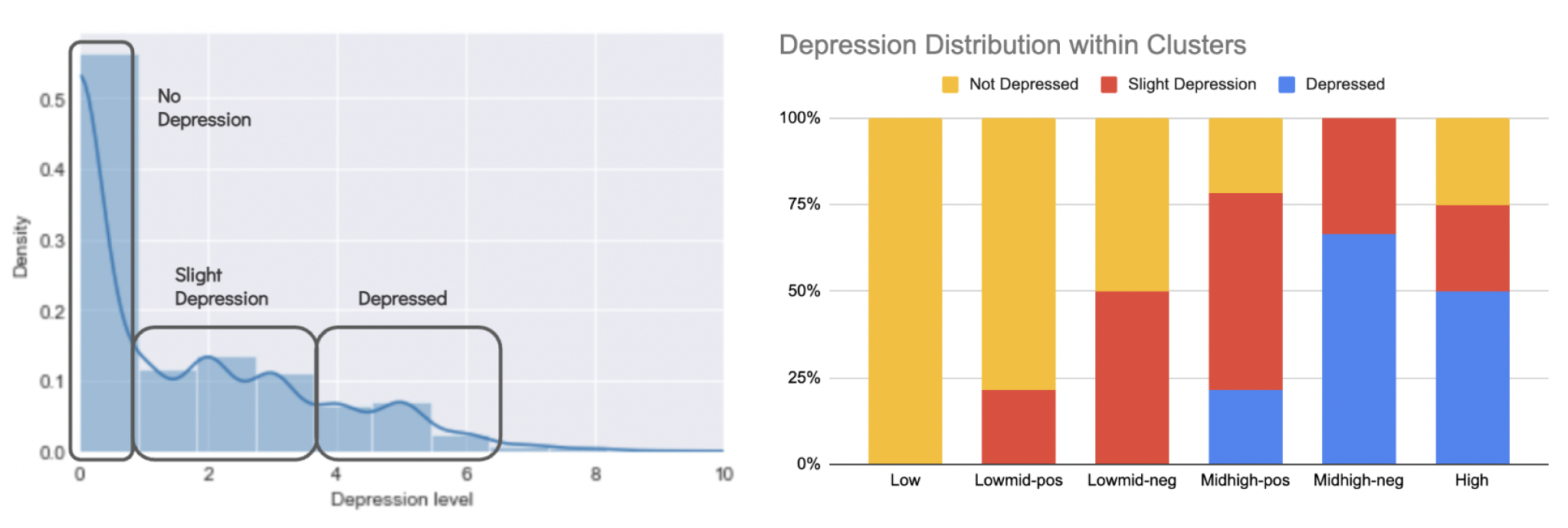
Right: Clustering based on the depression distribution. Left: Depression distribution within each fatigue cluster.

To dive more into the middle-high mesor with positive and negative acrophase clusters, we plotted each patient’s depression level throughout the day to a boxplot and find the distribution for patients with middle-high mesor and positive acrophase and patients with middle-high mesor and negative acrophase. We found that for middle-high mesor patients, acrophase might be able to determine the patient’s depression level (Figure 13).

**Figure 13.**
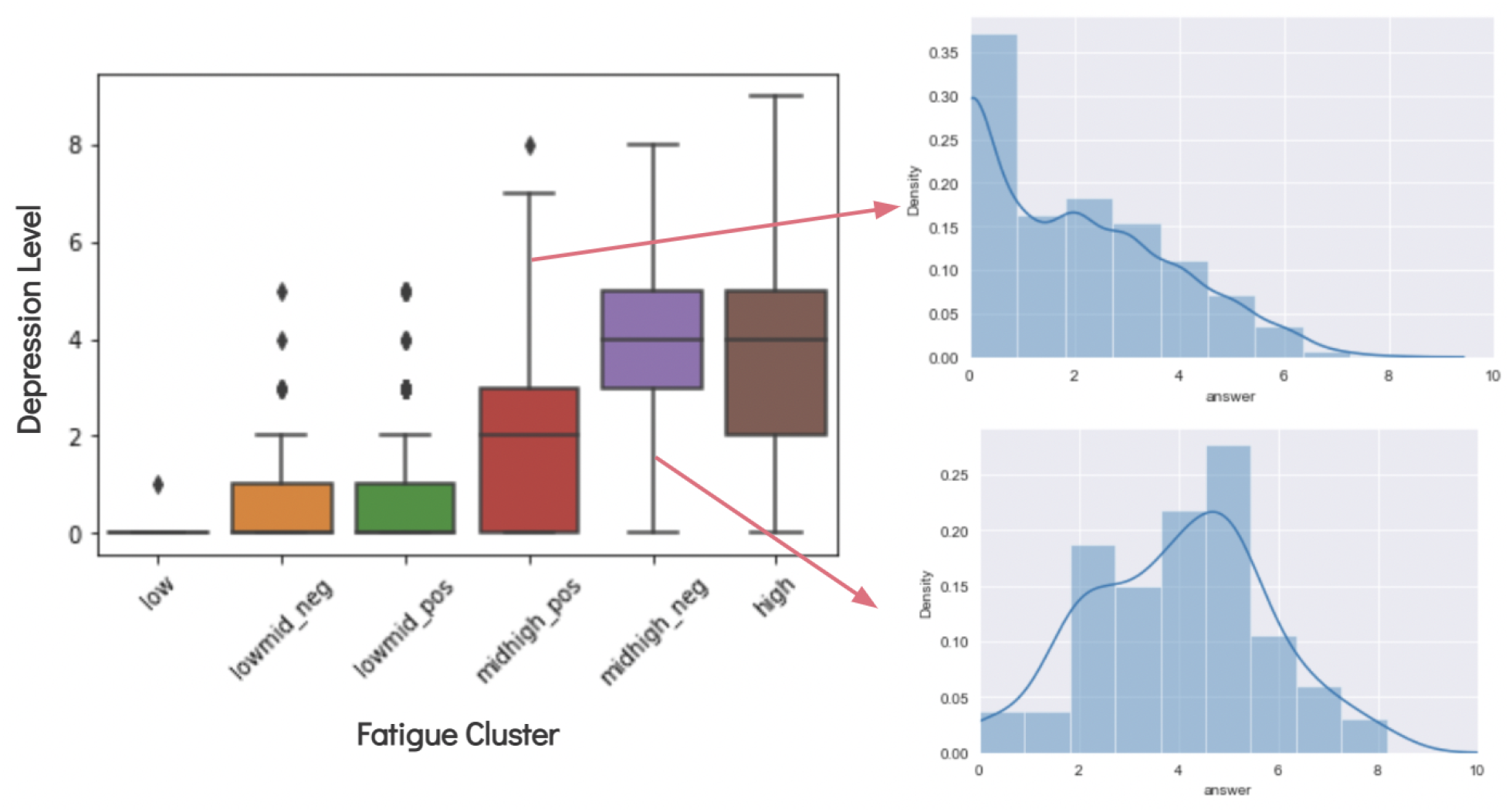
Left: boxplot of patients’ fatigue cluster compared to their depression level. Right: The distribution of depression for patients in middle-high mesor, positive acrophase and middle-high mesor, negative acrophase.

## 4. Discussion

This study is intended to be a pilot study for monitoring fatigue in multiple sclerosis patients. We have not separated patients with different medications, such as medication for anti-fatigue, anti-depressant, or anxiolytic medications, knowing that in the clinical routine, MS patients are often offered these drugs when there are fatigue and depression symptoms. There are also possibilities of many other confounding variables such as duration of MS, age, and sex that we have not taken into account in this study. We are also aware that most of our data population is people with white skin, therefore to generalize this study, we need more diverse data.

Many MS studies about anti-fatigue medication have not shown effectiveness in the treatment [14-17]. This might happen because of a difference in fatigue levels throughout the day and different circadian rhythms in each patient. Therefore, finding the stratification for patients with MS according to their fatigue phenotypes might be able to help future studies to find a personalized medication to alleviate fatigue in MS patients.

In this study, we interpolated our fatigue phenotype data with cosinor interpolation to mimic the circadian rhythm. Then we utilize dynamic time warping to find the similarities between each MS patient’s time series and then use hierarchical clustering to cluster the similarities between each patient. We also clustered the MS patients based on their cosinor parameters. After combining both clustering methods, we found 6 clusters, which are patients with low fatigue (low mesor), lower-middle mesor; positive acrophase, lower-middle mesor; negative acrophase, middle-high mesor; positive acrophase, middle-high mesor; negative acrophase, and high mesor. We also found an interesting relationship between the fatigue cluster with patients’ depression levels. We found that patients with middle-high mesor and positive acrophase are less likely to be depressed compared to middle-high mesor and negative acrophase.

We also have done some steps toward finding the association between fatigue clusters with their respective MRI images. We have developed an algorithm to anonymize personal health information from MRI images so that patients’ data can be protected.

### 4.1 Next Steps

In the future, we plan to continue the imaging work to find how different brain regions play roles in the stratified patients we have in this study. Finding the neurogenic mechanism of fatigue in MS patients will help to differentially assess specific treatments of fatigue in groups of MS patients with different fatigue phenotypes. Understanding patient stratification could provide MRI predictors of treatment response to support clinical decision-making by selecting the best fatigue management intervention for individual patients.

## Data Availability

All data produced in the present study are available upon reasonable request to the authors

https://brighammscenter.org/research/research-studies/climb-study

## Notes

### Competing Interest Statement

The authors have declared no competing interest.

### Clinical Protocols

https://brighammscenter.org/research/research-studies/climb-study

### Funding Statement

This study was funded by the Master's Thesis Program of the Master of Biomedical Informatics program at Harvard Medical School, in partnership with Brigham and Women's Hospital. No authors or their institutions received payment or services from a third party for any aspect of the submitted work.

### Author Declarations

The Institutional Review Board of Mass General Brigham gave ethical approval for this work (IRB Protocol #: 2017P001169), and informed consent was obtained from every study subject.

